# Successful contact tracing systems for COVID-19 rely on effective quarantine and isolation

**DOI:** 10.1101/2020.06.10.20125013

**Authors:** A. James, M.J. Plank, S. Hendy, R. Binny, A. Lustig, N. Steyn, A Nesdale, A Verrall

## Abstract

**Background:** Test, trace and isolate are the three crucial components of the response to COVID-19 identified by the World Health Organisation. Mathematical models of contact tracing often over-simplify the ability of traced contacts to quarantine or isolate.

**Method:** We use an age-structured branching process model of individual disease transmission combined with a detailed model of symptom onset, testing, contact quarantine and case isolation to model each aspect of the test, trace, isolate strategy. We estimated the effective reproduction number under a range of scenarios to understand the importance of each aspect of the system.

**Findings:** People’s ability to quarantine and isolate effectively is a crucial component of a successful contact tracing system. 80% of cases need to be quarantined or isolated within 4 days of quarantine or isolation of index case to be confident the contact tracing system is effective.

**Interpretation:** Provision of universal support systems to enable people to quarantine and isolate effectively, coupled with investment in trained public health professionals to undertake contact tracing, are crucial to success. We predict that a high-quality, rapid contact tracing system with strong support structures in place, combined with moderate social distancing measures, is required to contain the spread of COVID-19.

**Evidence before this study:** Existing models of contact tracing concentrate on the time taken to trace contacts and the proportion of contacts who are traced, often focussing on the differences between manual and digital tracing. They often over-simplify the quarantine and isolation aspect of contact tracing. For example, some models assume that isolation and quarantine are 100% effective in preventing further transmission, while others treat tracing coverage and isolation effectiveness as interchangeable. Numerous performance indicators have been used to measure the effectiveness of a contact tracing system. However, it is frequently not known how reliably these indicators measure the reduction in in onward transmission under a range of unknown parameters.

**Added value of this study:** We explicitly model the effectiveness of contact quarantine and case isolation in reducing onward transmission and show that these are not equivalent to tracing coverage. For example, isolating 50% of contacts with 100% effectiveness gives a much larger reduction in onward transmission than isolating all contacts but with only 50% effectiveness. We show that, although tracing speed is important, without effective isolation and quarantine it is a waste of effort. We show that seemingly straightforward indicators of contact tracing effectiveness are unreliable when the effectiveness of isolation is not guaranteed. We propose an indicator based on the time between quarantine or isolation of an index case and quarantine or isolation of secondary cases that is more robust to unknowns.

**Implications of all the available evidence:** Establishing support systems to enable individuals to quarantine and isolate effectively is equally important as implementing a fast and efficient contact tracing system. Effective contact tracing requires a skilled, professional workforce that can trace downstream contacts of a positive case, as well as upstream contacts to determine the source of infection and provide the high quality data needed. Over-reliance on digital contact tracing solutions or the use of untrained contact tracing staff are likely to lead to less favourable outcomes.

## Introduction

The WHO guidelines for control of COVID-19 emphasise three crucial components of an effective strategy: test, trace, and isolate ^1^. Collectively, this system of rapid case and contact management has become one of the key public health tools in the fight against COVID-19 worldwide ^2–4^. Contact tracing has been crucial in controlling several disease outbreaks, notably SARS, MERS and Ebola ^5,6^. While contact tracing alone is unlikely to contain the spread of COVID-19 ^7,8^, in countries like New Zealand, which is close to local elimination of the virus ^9^, it may allow population-wide social distancing measures to be relaxed. In countries with more widespread epidemics, it can allow safe reopening. There is a need for robust ways to measure and improve the effectiveness of contact tracing in reducing the spread of COVID-19. However, the effective reproduction number, *R_eff_*, is difficult to measure directly, is affected by numerous factors including population-wide restrictions, and can typically only be inferred after a substantial time lag ^10^ We therefore need reliable indicators for the effectiveness of contact tracing and other case-targeted interventions ^11^.

Existing mathematical models of contact tracing ^7,8^ focus almost exclusively on the time taken to trace contacts and the proportion of contacts who are traced. Most models assume that isolation is 100% effective in preventing further onward transmission from traced cases ^2^. Other models assume the effectiveness of isolation and the probability of being traced are interchangeable, i.e. a system where 50% of contacts are traced and isolation is 100% effective is assumed to have the same outcomes as one where 100% of contacts are traced and isolation reduces onward transmission by 50% ^7^. Quarantine refers to the separation of individuals who may have been exposed to the virus but are currently asymptomatic, and is distinct from isolation of symptomatic or confirmed cases ^12^. In reality, the quarantining of contacts and isolation of cases are complex and depend on a range of variables. Complete isolation of all confirmed cases is impractical in most countries: some contact with household members and essential service providers or healthcare workers is inevitable in at least some cases. Barriers to effective isolation are higher in communities with high levels of socioeconomic deprivation, insecure employment, and limited entitlement to paid sick leave. Quarantine of asymptomatic case contacts, the majority of whom are likely not infected, is even more challenging ^13^. In countries with large outbreaks, this could affect tens of thousands of people and require closure of entire workplaces for extended periods. As digital contact tracing systems are introduced, the number of false positives could increase further. Taking time off work to quarantine when asymptomatic is likely to be impossible for many. This suggests that quarantine is likely to involve precautionary measures rather than complete isolation and therefore to be less effective than isolation of confirmed cases.

We explicitly model the effectiveness of contact quarantine and case isolation in reducing onward transmission. We use a model calibrated using data on COVID-19 cases in New Zealand, where the virus has been controlled effectively and the number of current cases is very low. We also evaluate four potential performance indicators for the contact tracing system. We show that seemingly straightforward indicators, such as the proportion of cases quarantined before symptom onset, can be misleading. We propose an indicator based on the time between quarantine or isolation of an index case and quarantine or isolation of secondary cases. This may be harder to measure than some indicators, but is a more reliable measure of the reduction in the effective reproduction number. Our results highlight the importance of establishing support systems to enable individuals to quarantine and isolate effectively^14^. They also demonstrate that effective contact tracing requires a skilled, professional workforce that can trace downstream contacts of a positive case, as well as upstream contacts to determine the source of infection and provide the high quality data needed. This implies that over-reliance on digital contact tracing solutions or the use of contact tracing staff who are not trained in public health work are likely to lead to less favourable outcomes.

### Transmission, contact tracing, quarantine and isolation model

We use a continuous-time branching process model ^15^ to study the transmission of COVID-19 in the presence of contact tracing and case isolation (Figure 1) – see Supplementary Materials for details. Individual infectiousness is modelled using a time-dependent infection kernel connected with the time of symptom onset, with 35% of infections assumed to occur prior to onset ^16,17^. The key input parameters for the contact tracing model are: (i) the proportion of contacts successfully traced; (ii) the mean time taken to trace contacts following a positive test result in the index case; and (iii) the effectiveness of contact quarantine and case isolation in reducing onward transmission. The time for tracing ranges from zero, which can usually only be achieved for household contacts or by using a highly effective digital tracing system, through to 6 days on average. Traced contacts of a positive case who are not currently symptomatic go into home quarantine, i.e. minimise their interactions with others. Upon symptom onset, traced contacts isolated more stringently and tested for SARS-CoV-2. We assume that untraced symptomatic cases are also tested and isolated, but there is a delay from onset to isolation and testing (see Supplementary Materials for distributions). The model is based on New Zealand data which has a fast testing system with only one day on average between testing (when isolation starts) and result (when tracing starts). However, the shape of the assumed distribution for tracing time means the model can still be applied to countries where testing is slower by increasing the mean time from testing of the index case to tracing of contacts.

**Figure 1.**
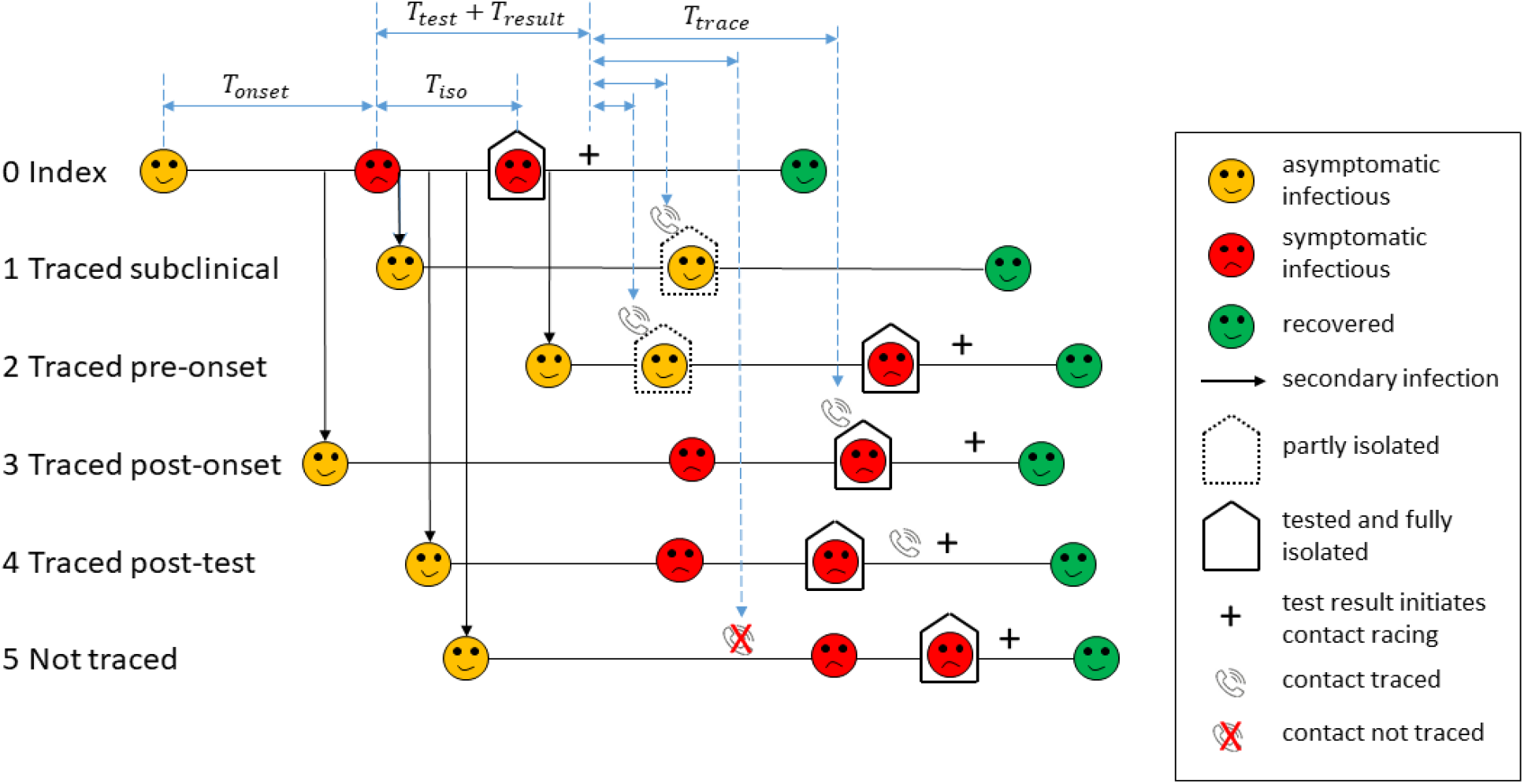
Schematic diagram of the contact tracing model. Infectious individuals are initially asymptomatic (yellow). For the index case who was not traced (0), there is a delay between onset of symptoms (red) and getting tested. Isolation occurs at some point between symptom onset and testing. There is a subsequent delay to the test result and tracing of contacts. Traced contacts (1-4) are quarantined when contacted by public health officials (phone icons) and isolated and tested immediately on symptom onset. Traced contacts (3) who are already symptomatic prior to being traced are isolated immediately when contacted. Traced contacts (4) that have already isolated prior to being traced are not affected. Contacts that cannot be traced (5) may still get tested and isolated, but this is likely to take longer. Asymptomatic individuals (1) do not get tested or isolated, but will be quarantined if they are a traced contact.

Measuring the input parameters empirically is not straightforward. In many countries, quarantine and isolation are left to the individual. Without strong government and community support, quarantine of asymptomatic or presymptomatic individuals in particular may be ineffective. Measuring the reduction in the number of onward transmissions during quarantine and isolation relies on high-quality data to compare contact rates of similar individuals across a broad sector of society. Even quantifying the proportion of contacts traced is not always straightforward due to imperfect recall.

### Determinants of the effectiveness of contact tracing

We run the model for three tracing probabilities, 0% (no contact tracing), 50% and 100% (all contacts traced), and a range of tracing speeds from instant tracing to 6 days on average. The reduction in onward transmission as a result of quarantine or isolation ranges from 50% to 100%. We assume that case isolation is always at least as effective as quarantine of asymptomatic contacts.

It is possible to have a case isolation policy with no contact tracing (Fig. 2, red lines). In this case, the more effective isolation is, the larger the decrease in *R_eff_* (compare Fig 2A, D, F, red lines). Across all isolation and quarantine scenarios, slow tracing results in poorer outcomes than fast tracing, i.e. higher *R_eff_* (Figure 2). If tracing is fast, then tracing more contacts reduces *R_eff_* further; if tracing is very slow (more than 5 days on average), the system is ineffective regardless of the proportion of contacts traced and the reduction in *R_eff_* is similar to the no-tracing or isolation only scenario.

**Figure 2:**
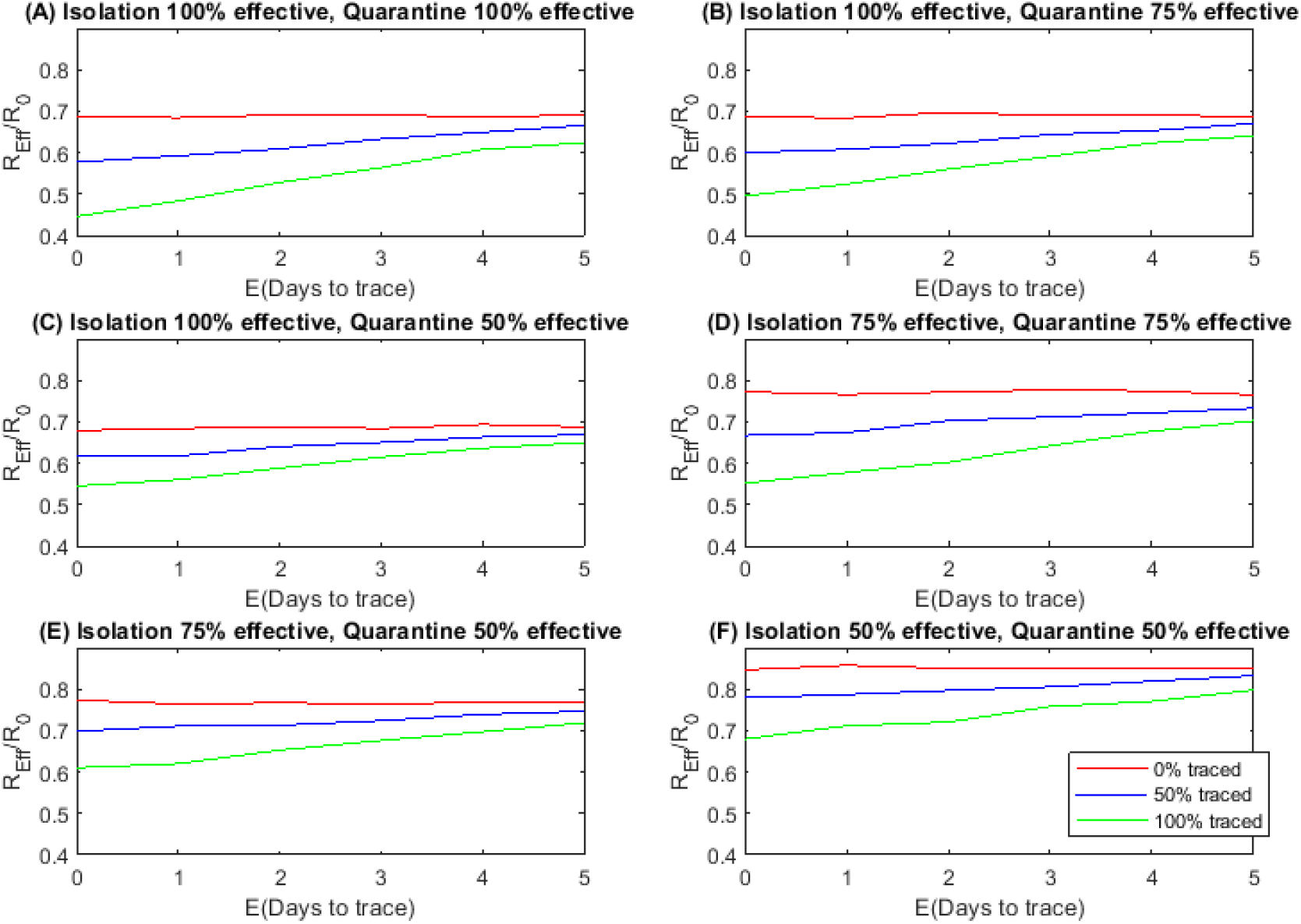
The effectiveness of a contact tracing and isolation system is strongly affected by the proportion of contacts traced, the tracing speed and the effectiveness of quarantine and isolation. Effectiveness measured by the reduction in effective reproduction number *R_eff_* relative to the no-control scenario against the expected number of days to trace contacts for a range of tracing probabilities and quarantine/isolation effectiveness.

In the results shown in Figure 2, we fixed the average time from testing to the test result being returned and varied the average time from a positive test result to being returned to tracing contacts. The key factor is the total delay from testing the index case to tracing contacts and the model is relatively insensitive to the breakdown of this total delay between testing and tracing. For example an average testing time of 1 day and an average tracing time of 5 days produces similar results to an average testing time of 3 days and an average tracing time of 3 days.

In reality, tracing speed and proportion of contacts traced may be linked: tracing a few contacts, particularly household or other close contacts, can be done quickly; but tracing a high proportion of all contacts will include contacts who are harder to trace, so the mean tracing time will increase. Our results show if the average tracing time is more than 6 days, there is little benefit trying to trace more contacts in terms of a reduction in *R_eff_* and the priority should be faster tracing.

The effectiveness of quarantine and isolation is a crucial determinant of the ability of the contact tracing system to reduce *R_eff_*. Because there is significant pre-symptomatic transmission of COVID-19 ^16,17^, fast tracing in conjunction with effective quarantine of contacts before symptom onset can greatly reduce spread. Although some countries, notably China, established arrangements for institutional isolation and monitored quarantine ^18^, most countries in Europe, North America and Australasia rely on home quarantine for contacts and home isolation for mild cases. Asking individuals to quarantine or isolate but failing to support them to do so means, for example, they will need to go shopping for food and other essential items, or have them delivered by family or friends who potentially should also be in isolation. Many individuals may be ineligible for paid sick leave, especially when asymptomatic. Crowded or unsuitable housing may mean isolation is not feasible. Precarious employment situations could be exacerbated by prolonged and possibly repeated absences. Given these realities, it is unlikely that 100% effective isolation and quarantine (Fig. 2A) is achievable. A more realistic scenario is where quarantine only reduces onward transmission by 50% (Fig. 2C). If tracing is instantaneous, this provides the same reduction in *R_eff_* as a mean tracing time of 2-3 days with 100% effective quarantine. If isolation of cases is only 50% effective (Fig. 2F), improving isolation may be more impactful than faster or more complete contact tracing (Fig. 2A, red line). Tracing 50% of contacts with 100% quarantine and isolation effectiveness (Fig 2A, blue line) is significantly more effective than tracing 100% of contacts with 50% isolation and quarantine effectiveness (Fig 2F, green line), whereas previous models ^7^ could not distinguish these two scenarios.

### Measuring the effectiveness of contact tracing

The key output of interest is the effective reproduction number *R_eff_*, defined in the model as the average number of secondary infections per case. It is difficult to measure *R_eff_* directly in practice because not all cases are detected and it is not always possible to link secondary cases with a unique index case. In addition, it is difficult to obtain reliable data to quantify the input parameters for the contact tracing model. The average tracing time is the easiest input parameter to measure, although this could be underestimated if some contacts present to healthcare before being traced. Officially reported values for the proportion of contacts traced typically represent the proportion of known contacts and do not include contacts that were not recalled by the case. Directly measuring the effectiveness of quarantine and isolation would require detailed information about the number of secondary infections during these periods. This would require intensive follow-up with quarantined and isolated individuals. Unless this was done for all cases, this intervention itself could bias the sample towards individuals with relatively effective isolation.

A robust performance indicator should: (i) be measurable from data routinely collected by public health organisations; and (ii) be closely correlated with the effective reproduction number *R_eff_* across a broad range of model inputs. In the absence of direct information about input parameters, we assessed the following potential performance indicators for the case isolation and contact tracing system^11^:

- Proportion of cases quarantined or isolated within 4 days of symptom onset in the index case.
- Proportion of cases quarantined or isolated within 4 days of quarantine or isolation of the index case.
- Proportion of cases with symptom onset within 4 days of symptom onset in the index case (i.e. serial interval less than 4 days).
- Proportion of cases quarantined before symptom onset.

The most robust indicator tested was the proportion of cases that were quarantined or isolated within 4 days of quarantine or isolation of the index case (Fig. 3A). This indicator is well correlated with *R_eff_* across variations in all three input parameters, meaning that improvements in contact tracing parameters are reliably associated with improvements in the indicator. In contrast, indicators that use onset time can be misleading. For example, the proportion of cases quarantined within 4 days of onset in the index case (Fig. 3B) depends almost exclusively on tracing speed and proportion of contacts traced, and is insensitive to the effectiveness of quarantine or isolation. If the effectiveness of quarantine and case isolation can be evaluated independently or assured in some other way, this could be a useful indicator of system effectiveness, but without this it is not useful. The serial interval (Fig. 3C) is determined predominantly by the virus transmission dynamics, in particular the incubation period. The effectiveness of quarantine has the strongest effect on this indicator by preventing onward transmission late in the infectious phase, but an improvement in either the tracing speed or the proportion of contacts traced leads to an apparent deterioration in the indicator. The reason for this counterintuitive result is that, as the contact tracing system becomes more effective in reducing transmission, the remaining cases contain a higher proportion of cases who were infected by subclinical carriers. These cases cannot be traced in the model and so continue to spread the virus relatively late in their infectious periods, leading to longer serial intervals.

**Figure 3:**
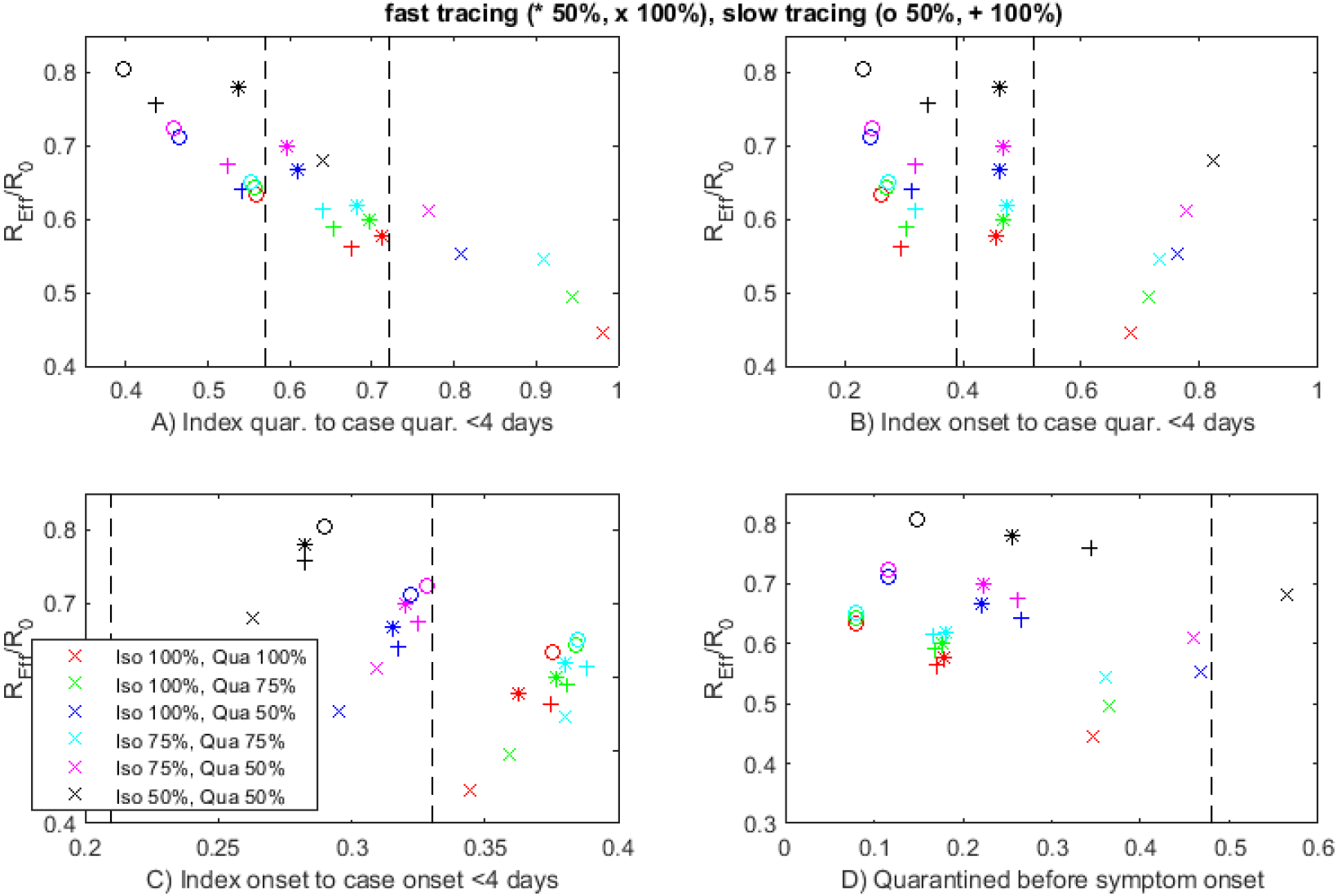
The proportion of cases quarantined or isolated within 4 days of the index case being quarantined or isolated (A) is the most robust indicator of the performance of the contact tracing system, measured by the reduction in effective reproduction number *R_eff_* relative to the no control case. Other indicators are not reliably correlated with *R_eff_* across all model parameters. Each plotted point corresponds to one combination of model parameters: fast tracing (same time as test result) of 50% of contacts (stars); fast tracing of 100% of contacts (crosses); slow tracing (on average 3 days after test result) of 50% of contacts (circles); slow tracing of 100% of contacts (pluses); varying effectiveness of isolation and quarantine are shown by different colours (see graph legend). Vertical dashed lines show the minimum and maximum values of the indicators computed from New Zealand case data.

The proportion of cases quarantined before symptom onset is the easiest indicator to measure as it can be calculated for all cases, including those that are not associated with a specific index case. However, it is the worst of the four indicators tested, showing apparently poorer outcomes as the effectiveness of quarantine or isolation improves (Fig. 3D). This happens because effective case-targeted interventions tend to prevent secondary infections that occur late in the infectious phase (e.g. case 2 in Fig. 1). The remaining cases are skewed towards those that were infected early in the infectious phase of the index case (e.g. case 3 in Fig. 1). These cases are the hardest to trace before symptom onset.

### Benchmarking against the New Zealand contact tracing system

We used the New Zealand EpiSurv dataset, centrally managed by ESR on behalf of the Ministry of Health (accessed 2 June 2020). We included the *N =* 95 cases with no recent history of overseas travel and with a symptom onset date between 8 April and 8 May. This minimises the impact of changes that may have occurred as a result of expansion of the contact tracing system at the start of the epidemic, and reduced contact rates following the introduction of strict social distancing restrictions on 26 March. Case discovery is categorised as either “contact of a case” (*N =* 82), “sought healthcare” (*N =* 9) or other. Of the 82 cases labelled as “contact of a case”, all were associated with at least one index case, allowing the values of the first three performance indicators defined above to be empirically calculated (see Supplementary Materials for details). The final indicator, proportion of contacts quarantined before symptom onset was calculated for all cases with available data.

Defining traced contacts to be the 82 cases labelled as “contact of a case” implies a tracing rate of 86%. Of these cases, 56% were quarantined or isolated prior to symptom onset. There was almost no difference in the average time from onset to testing between the traced and non-traced contacts (mean 2.8 days, standard deviation 2.6 days for non-contacts; mean 3.1 days, standard deviation 2.5 days for contacts). This suggests that many of the cases labelled as “contact of a case” may have self-identified or been identified *post hoc*, as opposed to being traced by public health officials. This reinforces the view that directly quantifying the proportion of contacts who are traced is difficult and that this may not be accurately reflected in routinely collected public health data.

To evaluate the effectiveness of the New Zealand contact tracing system, we calculated each of the four performance indicators across all cases in the sample (Fig. 3, vertical dashed lines). New Zealand Public Health Units have considerable contact tracing experience through routine management of tuberculosis cases, as well as previous epidemic outbreaks including measles and pandemic H1N1 influenza. Contact tracing staff have a background in public health and experience of managing the privacy issues that are involved in the work. Extensive interviews with cases establish potential source cases and identify contacts. Contacts are regularly monitored, often daily, to ensure adherence to quarantine restrictions, fast testing and an accurate onset time if symptoms develop. Quarantine and isolation are supported through workplace support and family care, and limited community quarantine or isolation facilities are available for those whose homes are unsuitable. The COVID-19 response was supplemented by a rapidly developed national contact management service to augment the scalability of local services and address fragmented information systems ^11^. A significant proportion of cases in New Zealand were associated with long-haul international travel, and tended to occur in relatively healthy, high socio-economic groups who were in a position to adhere to the restrictions. There were also cases in communities where financial and job insecurities were a significant issue. In addition, the data spans a period during which New Zealand was under the strictest social distancing measures, reducing contacts significantly ^19^. With no knowledge of the other system parameters using the recommended indicator (Fig. 3A) suggests that the contact tracing system reduced *R_Eff_* by 30-45% during this period. The noise in the data from self-reporting of isolation and symptom onset dates and the uncertainty in assigning index cases does not allow for a more accurate estimate. For comparison the indicator in Fig 3B, the proportion of cases where index onset to case quarantine is less than four days gives a much wider prediction of 20% to 45% *R_Eff_* reduction.

## Discussion and conclusions

Our results show that a high-quality, rapid contact tracing system, combined with strong support systems for people in quarantine or isolation, can reduce the effective reproduction number *R_eff_* by at most 60%. In the absence of any control interventions, the basic reproduction number *R*_0_ for COVID-19 is estimated to be between 2 and 4 ^20-22^. Containing the spread of COVID-19 requires *R_eff_* < 1, which implies that some level of moderate social distancing will likely be required during outbreaks in addition to case-targeted interventions. If case isolation or contact quarantine are imperfect, or some contacts are not traced or traced more slowly, then the reduction in *R_eff_* is only around 40%, meaning that stronger social distancing would be required to contain future outbreaks. In a country where testing is very slow, for example if it takes more than five days to return test results, it may be more effective to concentrate on highly effective isolation rather than contact tracing. Our model assumed that all clinical cases were diagnosed, which relies on widespread availability and uptake of testing. Case-targeted measures will be less effective if there is significant case under ascertainment.

We recommend using the time from quarantine or isolation of the index case to quarantine or isolation of secondary cases as the basis for measuring the performance of the contact tracing system. If at least 80% of cases are quarantined or isolated within 4 days of quarantining or isolating the index case, this indicates a reduction in #_e_// of at least 40%. For this to be effective, it has been suggested that the definition of a contact should be within 2 metres of an infected case for 15 minutes or more ^23^.

O’Dowd ^24^ and Verrall ^11^ identified key criteria against which to evaluate the contact tracing system, for example, suggesting that at least 80% of contacts must be quarantined within 4 days of symptom onset in the index case. We have shown that if the effectiveness of quarantine and isolation can be guaranteed, this criteria can be useful to benchmark a system. However, when these are not accurately known, the indicator recommended above is more robust. Case-targeted interventions tend to prevent onward transmission late in the infectious period. This skews remaining cases towards those infected in the pre-symptomatic or early symptomatic phase of the index case, and these are more difficult to trace in a timely way. Our work shows that in order to establish the effectiveness of a system, index-case pairs must be determined as accurately as possible, even if this is done *post hoc* with case investigation. Without this information, there is a danger that seemingly simple criteria, such as the proportion of cases quarantined before symptom onset, could give misleading indications of system performance.

The New Zealand contact tracing system is well-established and run by highly trained public health staff. Their work has been critical to the success of New Zealand’s elimination strategy for COVID-19 ^9^ However, even this well-established system suffers from noisy data and the difficulties of establishing index-case pairs. Our results show that even this success story can be improved, and, more usefully, point to the areas that need the most attention, in this case reducing the time taken to trace contacts. For countries with large outbreaks where contact tracing is either not well-established or does not have sufficient capacity to deal new daily cases, our results show that improving the effectiveness of case isolation may be better strategy initially.

Our model assumed that, in the absence of any case-targeted interventions, 35% of infections are subclinical with transmission rates reduced by 50% ^25^ and 35% of all onward transmission occurs during the pre-symptomatic phase. There is a wide range in empirically derived estimates for these parameters for COVID-19 ^2,16,17,26,27^, and the effectiveness of contact tracing is sensitive to these ^7^. If the true rates of asymptomatic and pre-symptomatic transmission are less than these assumed values, contact tracing will be more effective than our results predict. However, if the true values are higher, it is likely that contact tracing will need to be combined with stronger social distancing measures to contain COVID-19. In any case, the conclusion remains that the provision of systems to support people to quarantine and isolate effectively and the ability to rapidly trace the majority of contacts are crucially important. Our model does not distinguish between the different types of contact, such as household, work or casual, and each of these groups may have different tracing speeds and isolation effectiveness. This would be a useful addition to the model but would not undermine the conclusions on isolation effectiveness and robust indicators.

Further investment in improving the speed and capacity of contact tracing systems is needed. This is likely to be much more cost-effective than having to prolong or return to strict population-wide social distancing measures to contain a resurgence in cases. The crucial importance of support systems for people in quarantine or isolation and the ability to rapidly trace the majority of contacts mean that investment in skilled professionals and workers trained in public health work is essential. Overreliance on digital or automated contact tracing solutions or outsourcing case interviews and follow-up to undertrained staff are likely to substantially compromise these aspects of the system. Experience from the New Zealand contact tracing effort shows that the development of trusted relationships by public health officials and local community representatives is critical to an effective system. The success of contact tracing also relies on the universal provision of social security such as paid sick leave, leave entitlements for asymptomatic individuals in quarantine, and adequate job security and unemployment benefits.

## Data Availability

Data is available trough the New Zealand Ministry of Health. It cannot be made publicly available due to confidentiality concerns.

## Acknowledgements

The authors acknowledge the support of StatsNZ, ESR, the Ministry of Health and Barry Milne (Compass Research Centre, University of Auckland) in supplying data in support of this work. This work was funded by the Ministry of Business, Innovation and Employment, New Zealand and Te Pūnaha Matatini, the NZ Centre of Research Excellence for Complex Systems.

## Author contributions

AJ, MP developed the initial idea, and designed the methodology, AJ applied the model, NS, RB, AL curated data, SH provided project administration, All authors contributed to writing, review and editing.

## Data availability

For privacy reasons data is not publicly available. Please contact the authors or the New Zealand Ministry of Health to discuss availability for research purposes.

Supplementary Materials

### Contents

Materials and Methods: Model

Figure S1: Model generation time distribution.

Table S1: Parameter values and their sources.

## Materials and Methods: Model

We use an age-structured continuous-time branching process model ^15^ to study the transmission of COVID-19 in the presence of contact tracing and case isolation.

### Model assumptions

The key assumptions of the model are:

- Infected individuals fall into one of two categories: clinical or subclinical. Subclinical infections are assumed to have a reduced infectiousness (see Table S1 for parameter values). Note that subclinical individuals are asymptomatic during the entire infectious period and therefore are unlikely to be detected. Individuals who are pre-symptomatic are included in the clinical category.
- For clinical infections, the time between exposure and onset of symptoms (in days) *T_onset_* is drawn from a gamma distribution

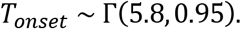
- The times of secondary infections *T_s_* are drawn from an individual-specific generation time distribution, which is shifted according to the time *T_onset_* of the index case ^7^ via:

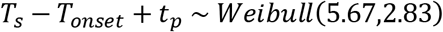

where *t_p_* > 0 is a constant chosen so that *p_pre_* = 35% of secondary infections occur prior to onset. Allowing for pre-symptomatic transmission in this way is realistic, but introduces the possibility that secondary infections could occur before infection of the index case. To prevent this, any randomly generated secondary infection times with *T_s_ < T_onset_* are discarded and another secondary infection time is generated instead. The population-wide generation time distribution is a model output (see Figure S1) and is consistent with empirical estimates ^2, 28, 29^.
- Symptomatic individuals who are not traced self-isolate after symptom onset. The time between symptom onset and isolation, *T_iSo_*, is Gamma distributed

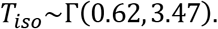 The distribution is found by fitting to the time between symptom onset and isolation in the NZ case data including all untraced individuals without a recent overseas arrival history and onset date between 8^th^ April and 8^th^ May.
- All clinical cases get tested. i.e. *p_test_ =* 1. In the absence of any contact tracing, the time, in days, between symptom onset and testing, *T_test_* is Gamma distributed

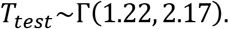 The distribution is found by fitting to the time between symptom onset and test date of the NZ case data of all untraced individuals without a recent overseas arrival history and onset date between 8^th^ April and 8^th^ May.
- Isolation reduces infectiousness to *c_iso_* of initial infectiousness; this is assumed to take effect from either the isolation date or the test date, whichever is earlier.
- The time from testing to test result, *T_result_*, (in days) is a minimum period plus an exponentially distributed random variable:

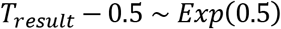 This is an approximate match to the New Zealand data.
- A positive test result initiates tracing of the index case’s contacts. It is assumed a proportion *p_trace_* of contacts are successfully traced, though cases may be symptomatic, have isolated and even been tested by the time they are traced.
- The time between the index case’s test result and tracing of contacts, *T_trace_*, is Gamma distributed with variance equal to 10% of the mean, i.e. tracing may be delayed but is completed quickly.
- Contacts traced before symptom onset go into partial isolation or quarantine, which reduces infectiousness to *c_quar_* of initial infectiousness. This is a weaker form of isolation during the pre-symptomatic stage (*c_quar_ ≥ c_iso_*).
- Once a traced contact develops symptoms, they are tested and fully isolated (infectiousness *c_iso_*) immediately (i.e. there is no delay from onset to testing or isolation). After the test-to-result delay described above, a positive test result initiates tracing of the secondary case’s contacts.
- Contacts traced after symptom onset immediately go into isolation and are tested.
- Subclinical individuals do not develop symptoms, do not get tested or isolated and their contacts are not traced.
- Subclinical individuals can be traced contacts of an index case and be quarantined. However, they do not develop symptoms and so will not get tested or fully isolated.
- If isolation is not 100% effective tracing continues during isolation and contacts are still traced with probability *p_trace_*

### Stochastic branching process model

The stochastic model for transmission of the virus is as follows:

- We segment the population into 3 age groups: 0-19 years, 19-65 years, over 65. The population is assumed to be well mixed within each group.
- The average reproduction number 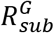 of subclinical individuals in any group *G* was assumed to be 50% of the average reproduction number 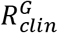 of clinical individuals in that group ^30^.
- In the absence of case isolation measures, each infected individual *i* causes a randomly generated number *N_i_~Poisson*(*R_i_*) of new infections.
- We assume a moderate level of social distancing which reduces transmission rates to 70%.
- As well as population heterogeneity across the different groups, individual heterogeneity in transmission rates was included by setting 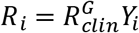 for clinical individuals in group *G* and 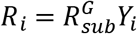 for subclinical individuals in group *G*, where *Y_i_* is a gamma distributed random variable with mean 1 and variance 2 ^31^.
- All individuals are assumed to be no longer infectious 30 days after being infected. This is an upper limit for computational convenience; in practice, individuals have very low infectiousness after about 14 days after symptom onset because of the shape of the generation time distribution (Fig. 1).
- The model is simulated using a time step of *δt =* 0.5 days. At each step, infectious individual *i* produces a Poisson distributed number of secondary infections with mean

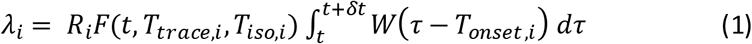

where *W* is the PDF of the Weibull distribution shown in Table S1, *R_i_* is individual *i’s* reproduction number, *T_trace_,_i_* is the time individual *i* was traced (if applicable), *T_iso,i_* is the time individual *i* was isolated (if applicable), and *F*(*t*) is a function describing the reduction in infectiousness due to isolation:

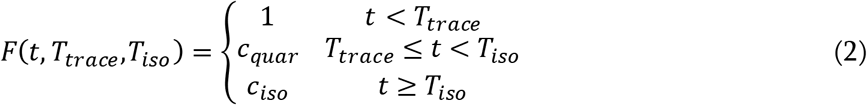
- The contact matrix Λ gives the probability Λ*_ij_* that a secondary infection originating from group *i* will be in group *j*, with *∑_j_*Λ*_ij_* = 1. New infections from group *i* are distributed across groups according to these probabilities.
- The contact matrix assumes 50% of contacts are from the same age group. The remaining 50% are distributed across all three age groups in proportion to their relative size in the population, consistent with ^32^.
- The model was initialised with 10 seed cases infected at time *t =* 0. *R_eff_* was calculated for each scenario using individuals who had experienced a full infectious period within 50 days of the seed cases.

### Performance indicators

For each combination of parameter values investigated, we calculated the values of the four following performance indicators:

1. Proportion of cases quarantined or isolated within 4 days of symptom onset in the index case.
2. Proportion of cases quarantined or isolated within 4 days of quarantine or isolation of the index case.
3. Proportion of cases with symptom onset within 4 days of symptom onset in the index case (i.e. serial interval less than 4 days).
4. Proportion of cases quarantined before symptom onset.

Time of quarantine or isolation means time of quarantine if there was a period of pre-symptomatic quarantine, or time of isolation otherwise. This is distinct from the time of diagnosis which may occur later for traced cases. These indicators were calculated in the model for all clinical cases (i.e. those that eventually became symptomatic), all of whom have an onset time and an isolation time. Asymptomatic cases were not included in the metrics, despite being possible for some cases, as these are unlikely to be available in collected data. This includes cases who were not traced before symptom onset, and whose isolation date is therefore after their onset date. Indicators 1-3 additionally require the index case to be clinical. Values of these indicators were averaged over 1000 simulations for each combination of parameter values investigated.

### Empirical calculation of performance indicators

We used the New Zealand EpiSurv dataset, centrally managed by ESR on behalf of the Ministry of Health (accessed 2 June 2020). We included the *N* = 95 cases with no recent history of overseas travel and with a symptom onset date between 8 April and 8 May. Case discovery is categorised as either “contact of a case” (*N* = 82), “sought healthcare” (*N* = 9) or other (*N* = 4). Of the 82 cases labelled as “contact of a case”, all were associated with at least one index case, allowing the values of the first three performance indicators defined above to be empirically calculated. For cases with multiple potential index cases, we calculated the following quantities

- Earliest time of onset in a potential index case 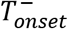.
- Latest time of onset in a potential index case 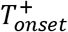.
- Earliest time of quarantine or isolation in a potential index case 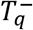.
- Latest time of quarantine or isolation in a potential index case 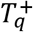.

Using 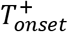 and 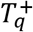 for all cases gave a minimum value for indicators 1-3. Using 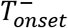 and 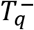 for all cases gave a maximum value for indicators 1-3. These minimum and maximum values corresponds to the two vertical dashed lines in Fig. 3A-C. Indicator 4 does not depend on the index case, so there is a unique value for this indicator (single vertical dashed line in Fig. 3D) which was calculated from all available case data.

**Figure S1.**
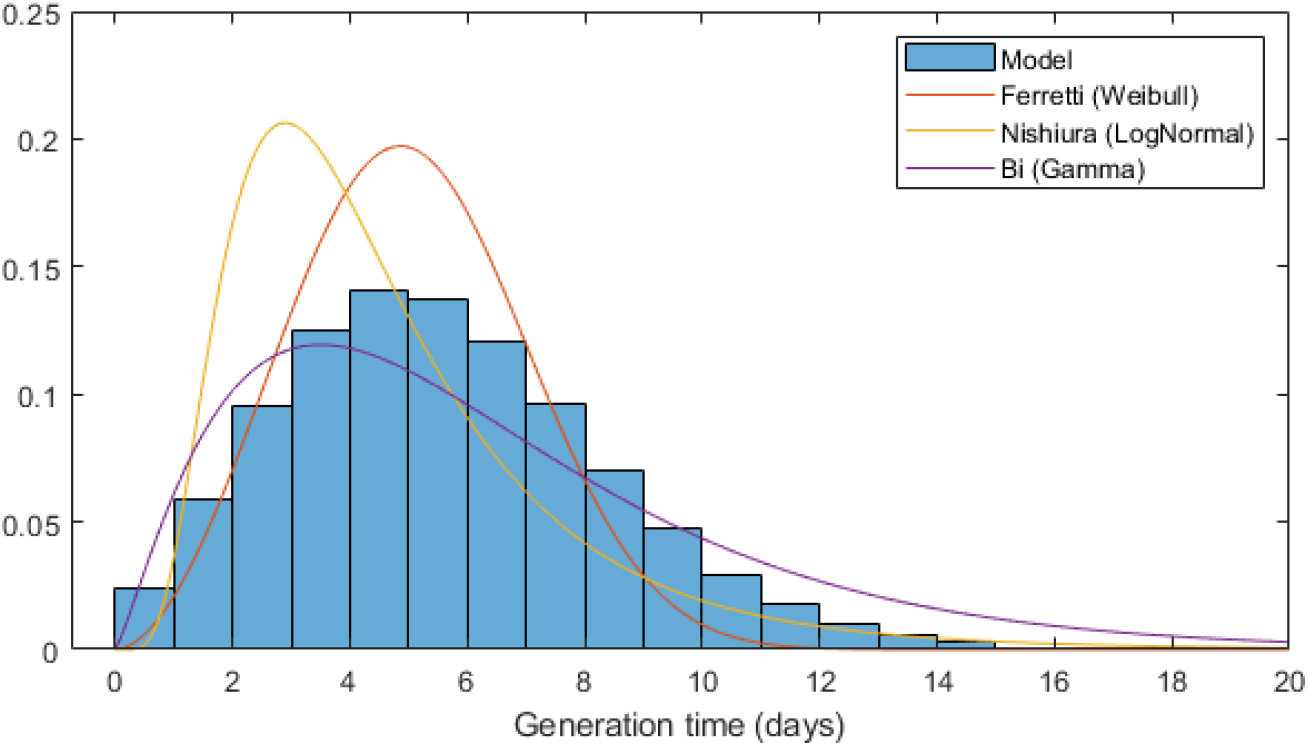
Distribution of generation times (time from infection of the index case to infection of secondary cases) with no contact tracing or case isolation. Three published generation time distributions are shown for comparison ^2,28,29^.

**Table S1.**
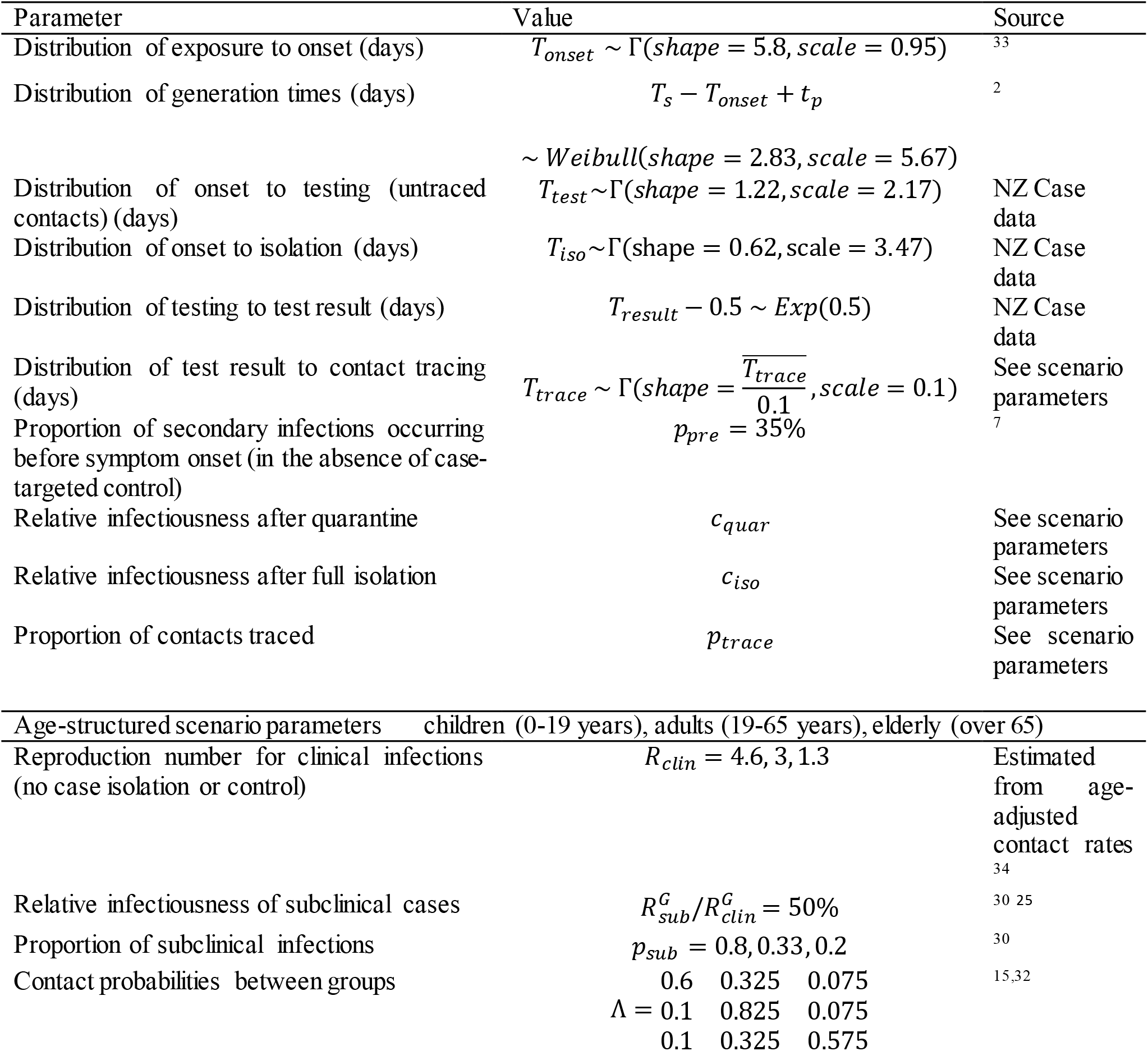
The parameters used in the model and their source.

## References

1. World Health Organization. Contact tracing in the context of COVID-19: interim guidance, 10 May 2020, 2020.

2. Ferretti L, Wymant C, Kendall M, et al. Quantifying SARS-CoV-2 transmission suggests epidemic control with digital contact tracing. Science 2020; 368(6491).

3. Ienca M, Vayena E. On the responsible use of digital data to tackle the COVID-19 pandemic. Nature medicine 2020; 26(4): 463-4.

4. Sun K, Viboud C. Impact of contact tracing on SARS-CoV-2 transmission. The Lancet Infectious Diseases 2020.

5. World Health Organization Centers for Disease Control Prevention. Implementation and management of contact tracing for Ebola virus disease: emergency guideline: World Health Organization, 2015.

6. Kang M, Song T, Zhong H, et al. Contact tracing for imported case of Middle East respiratory syndrome, China, 2015. Emerging infectious diseases 2016; 22(9): 1644.

7. Hellewell J, Abbott S, Gimma A, et al. Feasibility of controlling COVID-19 outbreaks by isolation of cases and contacts. The Lancet Global Health 2020.

8. Kucharski AJ, Klepac P, Conlan A, et al. Effectiveness of isolation, testing, contact tracing and physical distancing on reducing transmission of SARS-CoV-2 in different settings. medRxiv 2020.

9. Cousins S. New Zealand eliminates COVID-19. The Lancet 2020; 395(10235): 1474.

10. Obadia T, Haneef R, Boëlle P-Y. The R0 package: a toolbox to estimate reproduction numbers for epidemic outbreaks. BMC medical informatics and decision making 2012; 12(1): 147.

11. Verrall A. Rapid Audit of Contact Tracing for Covid-19 in New Zealand. Ministry of Health 2020.

12. World Health Organization. Considerations for quarantine of individuals in the context of containment for coronavirus disease (COVID-19): interim guidance, 19 March 2020: World Health Organization, 2020.

13. European Centre for Disease Prevention and Control. Contact tracing: Public health management of persons, including healthcare workers, having had contact with COVID-19 cases in the European Union - second update. Stockholm: ECDC, 2020.

14. Centre for Disease Control. Self-Isolation and Self-Quarantine Home Assessment Checklist for Coronavirus Dusease 2019 (COVID-19), 2020.

15. James A, Plank MJ, Binny RN, et al. A structured model for COVID-19 spread: modelling age and healthcare inequities. medRxiv 2020.

16. Ganyani T, Kremer C, Chen D, et al. Estimating the generation interval for coronavirus disease (COVID-19) based on symptom onset data, March 2020. Eurosurveillance 2020; 25(17): 2000257.

17. Tindale L, Coombe M, Stockdale JE, et al. Transmission interval estimates suggest pre-symptomatic spread of COVID-19. *MedRxiv* 2020.

18. Chen S, Zhang Z, Yang J, et al. Fangcang shelter hospitals: a novel concept for responding to public health emergencies. The Lancet 2020.

19. Binny RN, Hendy SC, James A, Lustig A, Plank MJ, Steyn N. Effect of Alert Level 4 on effective reproduction number: review of international COVID-19 cases. *medRxiv* 2020.

20. Liu Y, Gayle AA, Wilder-Smith A, Rocklöv J. The reproductive number of COVID-19 is higher compared to SARS coronavirus. Journal of travel medicine 2020.

21. Flaxman S, Mishra S, Gandy A, et al. Report 13: Estimating the number of infections and the impact of non-pharmaceutical interventions on COVID-19 in 11 European countries. 2020.

22. Jarvis CI, Van Zandvoort K, Gimma A, et al. Quantifying the impact of physical distance measures on the transmission of COVID-19 in the UK. BMC medicine 2020; 18: 1-10.

23. Keeling MJ, Hollingsworth TD, Read JM. The Efficacy of Contact Tracing for the Containment of the 2019 Novel Coronavirus (COVID-19). *medRxiv* 2020.

24. O’Dowd A. Covid-19: Johnson is on back foot over next steps to control pandemic. British Medical Journal 2020; 369.

25. Chau NVV, Lam VT, Dung NT, et al. The natural history and transmission potential of asymptomatic SARS-CoV-2 infection. *medRxiv* 2020.

26. Gudbjartsson DF, Helgason A, Jonsson H, et al. Spread of SARS-CoV-2 in the Icelandic population. New England Journal of Medicine 2020.

27. Lavezzo E, Franchin E, Ciavarella C, et al. Suppression of COVID-19 outbreak in the municipality of Vo, Italy. *medRxiv* 2020.

28. Nishiura H, Linton NM, Akhmetzhanov AR. Serial interval of novel coronavirus (COVID-19) infections. International journal of infectious diseases 2020.

29. Bi Q, Wu Y, Mei S, et al. Epidemiology and transmission of COVID-19 in 391 cases and 1286 of their close contacts in Shenzhen, China: a retrospective cohort study. The Lancet Infectious Diseases 2020.

30. Davies NG, Kucharski AJ, Eggo RM, Gimma A, Edmunds WJ, Group CC-W. The effect of non-pharmaceutical interventions on COVID-19 cases, deaths and demand for hospital services in the UK: a modelling study. MedRxiv 2020.

31. Lloyd-Smith JO, Schreiber SJ, Kopp PE, Getz WM. Superspreading and the effect of individual variation on disease emergence. Nature 2005; 438(7066): 355-9.

32. Prem K, Cook AR, Jit M. Projecting social contact matrices in 152 countries using contact surveys and demographic data. PLoS computational biology 2017; 13(9): e1005697.

33. Lauer SA, Grantz KH, Bi Q, et al. The incubation period of coronavirus disease 2019 (COVID-19) from publicly reported confirmed cases: estimation and application. Annals of internal medicine 2020; 172(9): 577–82.

34. Compass Research Centre. International Social Survey Programme (ISSP) 2017: Social Networks -New Zealand Survey. Auckland, New Zealand: University of Auckland; 2018.

